# Projecting the impact of SARS-CoV-2 variants on the COVID-19 epidemic and social restoration in the United States: a mathematical modelling study

**DOI:** 10.1101/2021.06.24.21259370

**Authors:** Rui Li, Yan Li, Zhuoru Zou, Yiming Liu, Xinghui Li, Guihua Zhuang, Mingwang Shen, Lei Zhang

## Abstract

**Background:** The SARS-CoV-2 Alpha variant B.1.1.7 became prevalent in the United States (US). We aimed to evaluate the impact of vaccination scale-up and potential reduction in the vaccination effectiveness on the COVID-19 epidemic and social restoration in the US.

**Methods:** We extended a published compartmental model and calibrated the model to the latest US COVID-19 data. We estimated the vaccine effectiveness against B.1.1.7 and evaluated the impact of a potential reduction in vaccine effectiveness on future epidemics. We projected the epidemic trends under different levels of social restoration.

**Results:** We estimated the overall existing vaccine effectiveness against B.1.1.7 to be 88.5% (95%CI: 87.4-89.5%) and vaccination coverage would reach 70% by the end of August, 2021. With this vaccine effectiveness and coverage, we anticipated 498,972 (109,998-885,947) cumulative infections and 15,443 (3,828-27,057) deaths nationwide over the next 12 months, of which 95.0% infections and 93.3% deaths were caused by B.1.1.7. Complete social restoration at 70% vaccination coverage would only slightly increase cumulative infections and deaths to 511,159 (110,578-911,740) and 15,739 (3,841-27,638), respectively. However, if the vaccine effectiveness were reduced to 75%, 50% or 25% due to new SARS-CoV-2 variants, we predicted 667,075 (130,682-1,203,468), 1.7m (0.2-3.2m), 19.0m (5.3-32.7m) new infections and 19,249 (4,281-34,217), 42,265 (5,081-79,448), 426,860 (117,229-736,490) cumulative deaths to occur over the next 12 months. Further, social restoration at a lower vaccination coverage would lead to even greater future outbreaks.

**Conclusion:** Current COVID-19 vaccines remain effective against the B.1.1.7 variant, and 70% vaccination coverage would be sufficient to restore social activities to a pre-pandemic level. Further reduction in vaccine effectiveness against SARS-CoV-2 variants would result in a potential surge of the epidemic in the future.

## Background

The COVID-19 pandemic remains a severe public health challenge despite the extensive public health interventions implemented worldwide. By 31^st^ May 2021, accumulatively 170.1 million infected cases and 3.6 million COVID-19-related deaths were reported worldwide [1]. Recent rollouts of effective COVID-19 vaccines have raised hope to control the pandemic. Countries leading the vaccination efforts have seen declining new infections and begun to relax in social distancing and travel bans. Experiences can be drawn from the United Kingdom (UK), Israel, and some states in the US (Vermont, Massachusetts, Hawaii, California), where vaccination coverage has reached 55-70% recently [2-4]. The vaccination program has successfully vaccinated more than 34 million people in the UK and prevented more than 10,000 deaths, enabling the government to ease previous tough restrictions [5]. Despite the promising progress, the goal of controlling or eliminating the COVID-19 epidemic is still far-reaching, primarily due to the insufficient vaccination coverage worldwide [6]. Besides, the emergence of new variants may hamper the effectiveness of the existing vaccines and shadow the potential population benefits of the vaccination.

Mutations in the receptor-binding domain (RBD) of SARS-CoV-2 enabled the virus to bind more effectively with the host receptor-Angiotensin converting enzyme 2 (ACE2) and enhanced better integration of the virus into the host [7]. The B.1.1.7 strain, with an N501Y substitution in RBD, has shown a 59% higher transmissibility and a 45% higher mortality rate compared to the wild type [8, 9]. Laboratory and clinical studies demonstrated that most COVID-19 vaccines remained effective against B.1.1.7 [10-13]. However, the additional E484K substitution of RBD in B.1.1.7 and the emergence of more transmissible Delta variant B.1.167.2 cast doubts on the effectiveness of the existing vaccines [14-16].

The US has experienced multiple waves of severe COVID-19 epidemics in the past [1]. In response, the US has developed effective vaccines against COVID-19 [17-19]. The US has seen a rapid rollout of a comprehensive vaccination program since 13^th^ December 2020 [20]. By 31^st^ May 2021, the vaccination coverage in the US population has reached 51% [21]. The US has established a comprehensive surveillance system that closely tracked the spread of the COVID-19 variants [22]. The latest statistics reported that 70% of COVID-19 diagnoses in the US belong to the B.1.1.7 variant [23]. Thus, the US provides an ideal setting for evaluating the benefits of mass vaccination for the COVID-19 epidemics with the emergence of new variants.

Numerous modelling studies have simulated vaccination impact on COVID-19 in the US. Most of these studies have consistently demonstrated that effective vaccination would significantly reduce new infections and among the infected, the clinical adversities, ICU admissions and mortality [24-26]. Studies also indicated that a population vaccination coverage of at least 50-80% is required to reduce the effective reproductive number to below one and enables restoration of social activities to a pre-pandemic level [27, 28]. Some studies have evaluated the transmission of hypothetical SARS-CoV-2 variants before the actual report of B.1.1.7 in the US [29, 30]. They have substantially underestimated either the transmissibility of B.1.1.7 or the update of vaccination in the US in these models, leading to a biased projection of the epidemic trends. A more precise evaluation of the impact of vaccination and new variants is warranted to inform the future epidemic and relevant public health interventions.

Our study aims to project the epidemic trend of the COVID-19 epidemic amid increasing vaccination coverage in the US. We further evaluate the potential population impact on the epidemic trends if further new variants emerge with higher transmissibility, mortality, and lower vaccine effectiveness. Our findings will help inform public health measures for epidemic management of new variants in the US.

## Methods

### Data sources

We collected publicly available reportable epidemiological data in the US from Johns Hopkins University Coronavirus Resource Center [1] and the Centers for Disease Control and Prevention (CDC) [21, 23]. These websites provided daily confirmed COVID-19 infection cases and death cases from 1^st^ March 2020 to 31^st^ May 2021, and the number of COVID-19 vaccine uptakes and variant proportions from 13^th^ December 2020 to 31^st^ May 2021. All four types of data were used to calibrate the model (details in Supplementary Materials).

### Model structure

We extended a previously published dynamic compartmental model [27] to describe the circulation of two SARS-CoV-2 strains (the wild type and B.1.1.7 variant) in the US. Our model consisted of eighteen compartments (Figure S1). A susceptible or vaccinated individual (S, V) may be infected by a SARS-CoV-2 strain (either the wild type or B.1.1.7) and entered a latent infection stage (E, E_m_, *m* denotes B.1.1.7 variant). After a mean incubation period of 5.2 (4.1-7.0) days [31], a proportion of infected individuals developed symptoms (I_1_, I_m,1_) before being diagnosed and documented (T_1_, T_m,1_). The remaining asymptomatic infections (A, A_m_) would spontaneously recover (R, R_m_). Undiagnosed and diagnosed infected individuals may progress to severe/critical stage (I_2_, I_m,2_, T_2_, T_m,2_) and die (D, D_m_) or recover (R, R_m_). The description of progression rates, derivation of the force of infection and mathematical expressions were documented in the Supplementary Materials.

### Model assumptions

We assumed that B.1.1.7 was 59% (56-63%) more transmissible and 45% (18-78%) more deadly than the wild type SARS-CoV-2 [8, 9]. We also assumed the recovered individuals could not be reinfected by any strains of SARS-CoV-2, suggesting a complete cross-protection and no co-infection of SARS-CoV-2 strains. In this study, we calculated the weighted-average effectiveness (91.4%) of the three available vaccines, Pfizer (92.6%), Moderna (92.1%), and J&J Jensen (66.9%), based on their population coverage in the US [17-19, 32]. We assumed the protection of natural and vaccine-induced immunity was life-long.

### Model calibration

We calibrated the model using a nonlinear least-squares method that minimised the Root Mean Squared Error (RMSE) between model-simulated and reported data. Based on the ‘calibrated’ scenario, we perturbed model parameters around the ‘calibrated’ parameter set to generate a band of curves that best describe the data variations and retain a minimal level of RMSE. We randomly generated 200 small ‘perturbating factors’. For each of the perturbing factors, we randomly sampled 100 parameter sets based on Latin Hypercube Sampling (LHS) between the parameter range generated by a random walk (adding the perturbing factor in both positive and negative directions). Hence, we obtained 200 groups of various perturbance, and each group has 100 randomised parameter sets. For each of the 200 groups, we calculated the number of data points covered by the band of curves simulated by the 100 parameter sets and their RMSE. We hence selected the one with the minimal RMSE across 200 bands as the set of simulations that best explained the observed data. The 100 curves in the selected band were used to calculate the 95% CI of the model outcomes. The vaccine effectiveness against B.1.1.7 was estimated spontaneously during model calibration. We validated that the estimated effectiveness of 88.5% produced the lowest RMSE in Figure S2.

### Impact of reduction in vaccine effectiveness

The emergence of E484K substitution in B.1.1.7 or other new variants may potentially reduce the effectiveness of the existing vaccines. We evaluate its impact on the COVID-19 epidemic when the vaccine effectiveness (1) adopts the model-estimated value for B.1.1.7 (baseline scenario); reduces to (2) 75%; (3) 50% and (4) 25%. For each scenario, we calculated the cumulative infections and deaths due to COVID-19 over the next 12 months (1^st^ June 2021 to 31^st^ May 2022). Since we did not know the viral properties of potential new variants, we also simulated nine scenarios with varying viral transmissibility and mortality as a sensitivity analysis (Figure 2).

### Impact of social restoration

Social restoration would significantly enlarge the force of infection for both wild type and B.1.1.7 variant. To assess the timing of social restoration and herd immunity, we conducted three scenarios of social restoration at 60%, 65%, and 70% vaccination coverage levels and calculated the daily and cumulative COVID-19 infections and deaths over the next 12 months (Figure 3).

### Uncertainty and sensitivity analyses

Based on the selected 100 parameter sets in model calibration, we produced the sensitivity analysis to accommodate the uncertainty of model parameters and determine the 95% CI of the cumulative COVID-19 infections and deaths. In addition, we also explore the impact on the epidemic trends of COVID-19 in several scenarios (new variants emerge with higher transmissibility, mortality, and lower vaccine effectiveness; social restoration; Figures 2-3). All analyses and simulations were performed in MATLAB R 2019b.

## Results

### Model estimated slightly reduced vaccine effectiveness against B.1.1.7

Our calibrated model estimated the overall existing vaccine effectiveness against B.1.1.7 to be 88.5% (95%CI: 87.4-89.5%) (Figure 1, additional validation in Figure S2), slightly lower than the weighted average effectiveness (91.4%) among the three vaccines available in the US.

**Figure 1.**
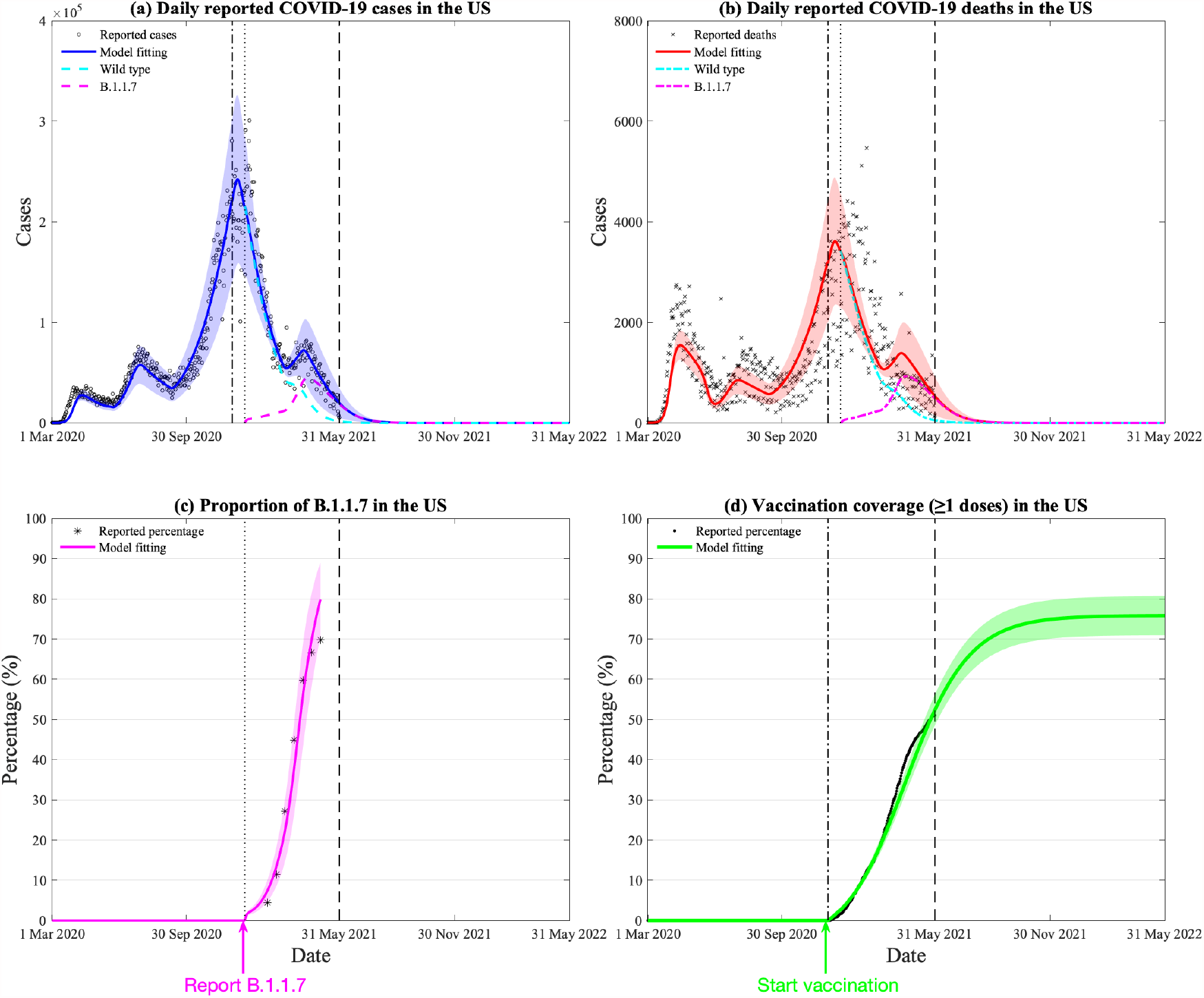
Model calibration and estimation of vaccine effectiveness on B.1.1.7 variant by daily COVID-19 cases, deaths, vaccination, and proportion of B.1.1.7 in the US. (a) Model calibration based on 15-month daily reported COVID-19 cases in the US, the blue area denotes the 95% confidence interval, the black dash-dotted and dotted lines denote the beginning date of vaccination and reporting B.1.1.7 variants, the green and cyan dashed lines denote the daily reported cases of wild type and B.1.1.7. (b) Model calibration based on 15-month daily reported COVID-19 deaths in the US. (c) Reported and model-fitted proportion of B.1.1.7 (of all reported cases) in the US. (d) Reported and model-fitted vaccination coverage (≥1 doses) in the US.

**Figure 2.**
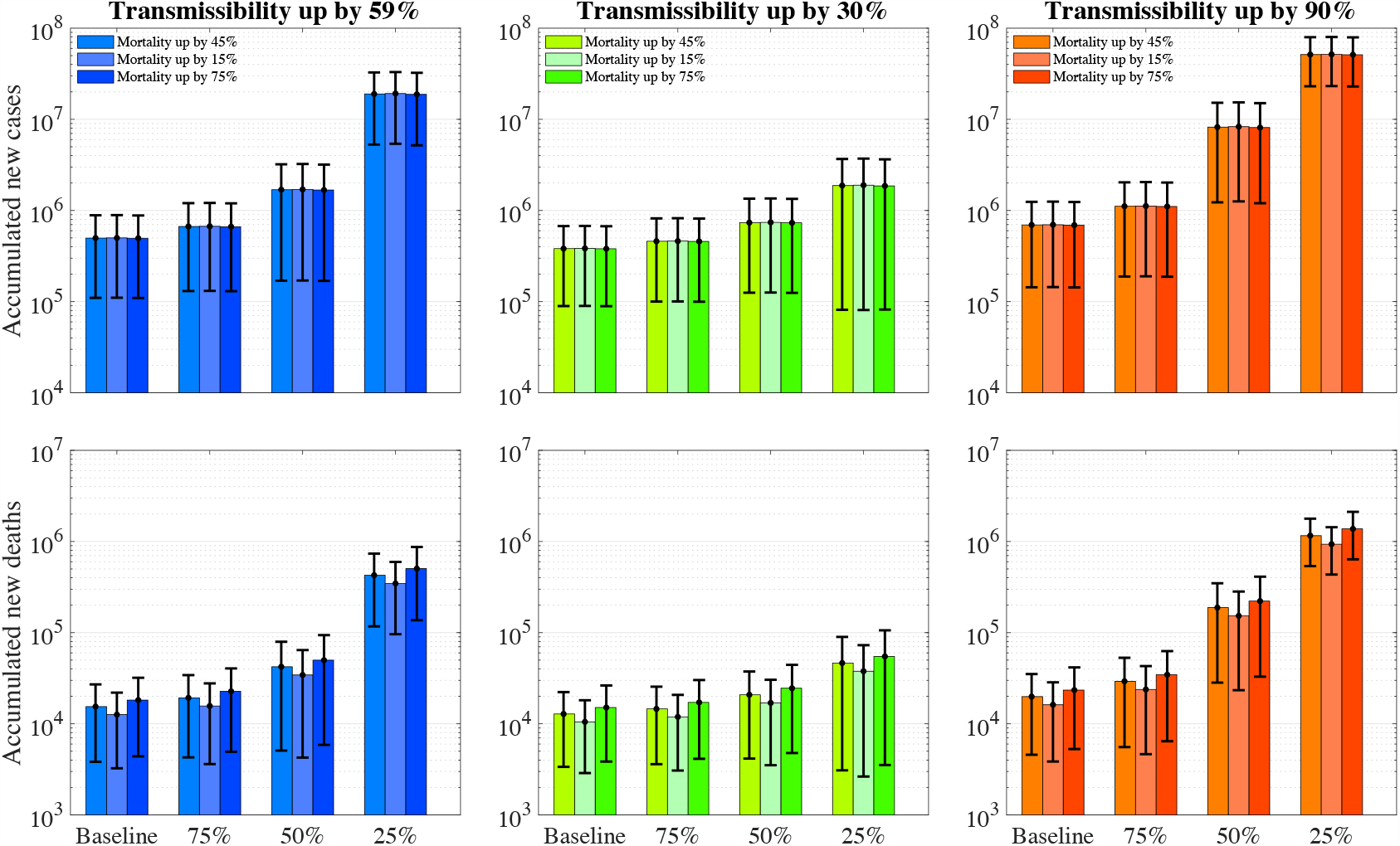
The number of accumulated new COVID-19 infections and deaths over the next 12 months with varying vaccine effectiveness, viral transmissibility and mortality. The first row represents the accumulated number of new infections whereas the second row represents accumulated deaths. The colour (blue, green and orange) bars denote the varying transmissibility of SARS-CoV-2 variants. The error bars represent 95% confidence intervals.

**Figure 3.**
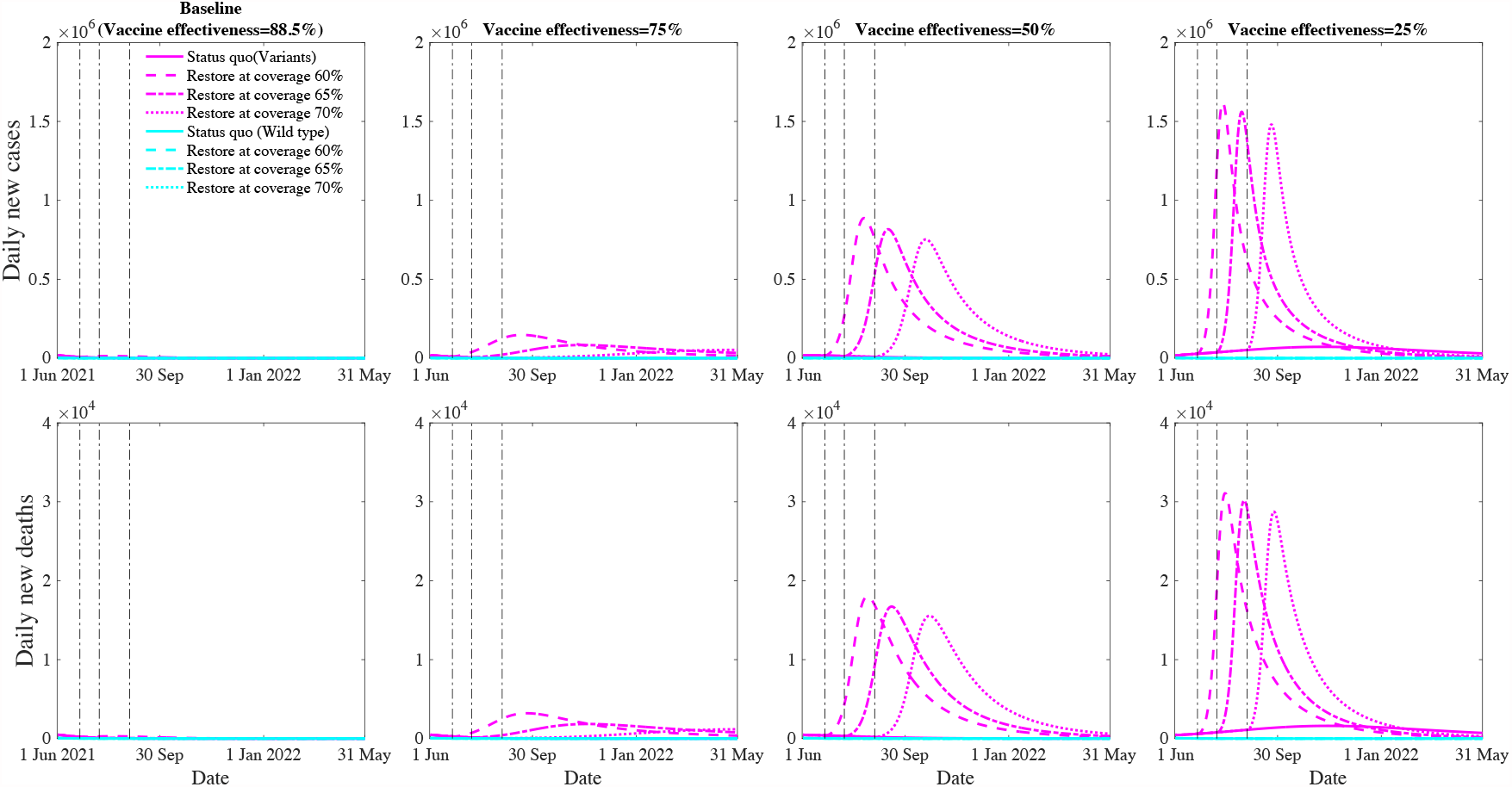
Projected COVID-19 new cases and deaths with social restoration at vaccination coverage of 60%, 65%, and 70% level over the next 12 months. The first row represents the daily number of new infections whereas the second row represents daily number of deaths. The columns represent an overall vaccine effectiveness of 88.5% (estimated effectiveness for B.1.1.7), 75%, 50% and 25%, respectively. The magenta and cyan lines (solid, dashed, dash-doted, dotted) denote various restoration scenarios for the wild type and COVID-19 variants.

### B.1.1.7 would dominate over the existing COVID-19 wild type strain

Our model demonstrated that the estimated B.1.1.7 trend closely resembled the reported data (Figure 1c). Since March 2021, B.1.1.7 had quickly expanded and dominated other SARS-CoV-2 lineages. We predicted that B.1.1.7 would continue to increase if no new variants emerge.

### Vaccination would control the COVID-19 epidemic with persistent public health interventions

Despite the rise of B.1.1.7, our model predicted that the increasing vaccination and persistent public health interventions would control the COVID-19 epidemic over the next 12 months (1^st^ June 2021 to 31^st^ May 2022). If the current trend of vaccination is sustained, vaccination coverage (having received ≥1 doses) in the US would exceed 70% by the end of August 2021 and a plateau around 75% at the end of November 2021 (Figure 1d). Assuming all interventions remained at the current levels, we anticipated 498,972 (109,998-885,947) cumulative infections and 15,443 (3,828-27,057) cumulative deaths nationwide over the next 12 months. Of which, 95.0% (81.8-96.7%) infections and 93.3% (76.7%-95.6%) deaths were caused by B.1.1.7.

### Potential population impact of vaccine effectiveness reduction of a new variant

We simulated the potential impact of vaccine effectiveness reduction on the epidemic trend due to the emergence of new SARS-CoV-2 variants (e.g., E484K substitution in a B.1.1.7; Delta variant [14, 16]). If the new variant would reduce vaccine effectiveness to 75% yet retain similar transmissibility and mortality as B.1.1.7, estimated 667,075 (130,682-1,203,468) cumulative infections and 19,249 (4,281-34,217) cumulative deaths over the next 12 months. Likewise, if vaccine effectiveness would reduce to 50% and 25%, there will be 1.7m (0.2-3.2m), 19.0m (5.3-32.7m) cumulative infections and 42,265 (5,081-79,448), 426,860 (117,229-736,490) cumulative deaths over the next 12 months, respectively.

### Potential population impact of transmissibility of a new variant

The emergence of a more transmissible COVD-19 variant (e.g. B.1.167.2, up by 90% of the wild type) than the current B.1.1.7 strain (up by 59% of the wild type) but share a similar mortality rate and vaccine effectiveness would result in 694,193 (143,823-1,244,563) cumulative infections and 19,861 (4,579-35,143) cumulative deaths over the next 12 months (Figure 2). In contrast, if its transmissibility is only half of the B.1.1.7 strain (up by 30% of the wild type), the estimated number of cumulative infections and deaths would only be 381,684 (89,438-673,930) and 12,824 (3,371-22,276) over the next 12 months. However, we acknowledge the latter scenario may not happen due to the less competitive nature of the variant.

### Potential population impact of the mortality of a new variant

The emergence of a more deadly COVD-19 variant (up by 75% of the wild type) than the current B.1.1.7 strain (up by 45% of the wild type) but share a similar transmissibility rate and vaccine effectiveness would result in more deaths but fewer infections, estimated 495,830 (109,604-882,056) cumulative infections and 18,197 (4,393-32,002) cumulative deaths over the next 12 months (Figure 2). In contrast, a reduced mortality (up by 15% of the wild type) would only cause a similar number of new infections and less deaths (12,615 [3,250-21,980]).

### Potential population impact of social restoration at the various vaccination coverage level

Retaining high vaccine effectiveness enables a sooner social restoration (Figure 3). We projected that, with the estimated effectiveness (88.5% against B.1.1.7), restoring social activity to the pre-pandemic level at the 60%, 65% and 70% vaccination coverage would result in 1.6m (0.2-2.9m), 0.7m (0.1-1.2m), 511,159 (110,578-911,740) cumulative infections and 39,040 (5,509-72,570), 19,562 (3,873-35,250), 15,739 (3,841-27,638) cumulative deaths over the next 12 months, respectively. However, if the vaccine effectiveness dropped to 75%, social restoration at the 60% vaccination coverage would result in a significant further wave that causes 22.7m (14.2-31.3m) cumulative infections and 513,154 (320,260-706,048) cumulative deaths. Further, if the vaccine effectiveness dropped to 25%, social restoration will not be possible, and current public health interventions need to be further strengthened.

## Discussion

Our study assesses the potential impact of vaccination on the COVID-19 epidemic over the next 12 months, with the consideration of an increasing spread of B.1.1.7 and other potential variants. We identified several key findings. First, our model estimated the current vaccine effectiveness against B.1.1.7 is about 88.5%. With this effectiveness, we project that the number of new infections would be depleted by 98% over the next 3 months, as the population vaccination coverage increases to 70% during the same period. This enables potential social restoration to the pre-pandemic level. However, if the vaccine effectiveness against B.1.1.7 is reduced to 25% due to the E484K mutation, our model predicted a further wave of the epidemic may be inevitable. Further, if a 30% more transmissible variant emerges, then vaccine effectiveness needs to be above 50% to prevent a major resurge of the epidemic.

Our finding demonstrates the overall vaccine effectiveness for B.1.1.7 is slightly lower than that for the wild type. This is consistent with previous findings. Laboratory studies based on serum and plasma neutralisation assay with authentic and pseudoviruses of B.1.1.7 show no significant change in neutralising activity of current vaccines against B.1.1.7, despite the presence of RBD mutations [11, 12, 14, 33-36]. However, Emary, al et. [10] find *in vitro* that neutralising antibody titres generated by AstraZeneca are lower for B.1.1.7, but the clinical vaccine effectiveness remains at 70·4%, indicating a lower activity may still be sufficient to confer protection or elicit host immunity. Further studies provide evidence of vaccine protective effects for B.1.1.7 in a population [13, 37-40]. Abu-Raddad, al et. [37] reported an estimate of 87.0% effectiveness of Pfizer against the B.1.1.7 variant. Consistently, Haas, al et. [38] reported an overall effectiveness of Pfizer vaccine to be 95.3% in Israel, where the epidemic is dominated by B.1.17 (94.5%). Lopez Bernal al et. [40] estimated a combined 60% vaccine effectiveness for both Pfizer and AstraZeneca in England, where B.1.1.7 is first reported. Nevertheless, with this high effectiveness for B.1.1.7, our study supports that current vaccination effort will control the COVID-19 epidemic in the US.

Our finding also suggests that a new variant (e.g., Delta) or an E484K-mutated B.1.1.7 variant may reduce the overall vaccine effectiveness and become dominant in the US. In this case, we project further waves of the epidemic with its size proportional to the level of vaccine effectiveness reduced. To cater for these scenarios, we recommend several potential strategies. First, maintaining the current vaccination effort is of vital importance for COVID-19 control even with reduced vaccine effectiveness. This is because a weakened vaccine still reduces the overall number of new infections and the risk of new variants. Besides, even though a vaccine may not completely protect against the infection, it still significantly reduces the clinical severities of the infected individuals [18, 32]. Second, the pharmaceutical industry should prioritise its efforts in developing new vaccine boosters for the emerging new variants of concerns [41, 42]. At a population level, epidemiological studies need to focus on whether a booster shot, or third dose of the existing vaccines, or a combination of both should be implemented to maximise vaccination protection. Third, orchestrated vaccination efforts worldwide are necessary to ensure less-developed regions can receive sufficient vaccines for epidemic control and hence reduce the risk of new COVID-19 variants.

Our finding indicated that social restoration is strongly dependent not only on the vaccine effectiveness but also on its coverage. We discovered that 70% vaccination coverage with the current vaccine effectiveness might allow safe restoration of social activities to the pre-pandemic level. However, the decline of vaccine effectiveness would lead to further waves of epidemic and reduce the likelihood of social restoration. In fact, if the vaccine effectiveness reduces to 25%, current public health interventions in the US would need to be further strengthened to control the epidemic. As the widespread SARS-CoV-2 would result in a higher likelihood of variants and reduce vaccine effectiveness, which in turn causes further spread of the virus [7, 43, 44], a high vaccination coverage will limit the viral spread and break the vicious cycle. Therefore, encouraging the community to vaccinate is essential to reduce variants and retain vaccine effectiveness for social restoration. Authorities should enhance health promotion to reduce the public’s misunderstanding about vaccination and provide free and accessible vaccination [45]. Besides, relaxing restrictions for fully vaccinated individuals in public places and travelling is also an alternative to encourage the community to vaccinate.

This study has several limitations. First, our model did not consider age structure and variations in the risk of infection and mortality across age groups. Second, our model structure only investigated one SARS-CoV-2 variant. We have mostly focused on B.1.1.7 since B.1.1.7 is the most reported variant in the US but only included other variants as a part of sensitivity analysis. The latest report has indicated that Beta (B.1.351), Gamma (P.1) variants and Delta (B.1.617.2) have accounted for 11.9% of new infections, and the proportion is growing [23]. Modelling the competition of variants would be our future investigation. Third, we assumed that the natural immunity and immunity elicited by vaccines was life-long. If the neutralising antibody reduction results in reduced vaccine protection, more strict public health interventions or a new vaccine booster for enhancing the immune response may be necessary. Finally, the model did not consider the impact of the individual willingness of vaccination on herd immunity.

## Conclusions

In conclusion, our modelling exercise indicates that the current vaccines in the US remain effective for B.1.1.7, and 70% of vaccination coverage would be sufficient to restore social activities to a pre-pandemic level if no new, more transmissible variants emerge. The emergence of new, more transmissible variants accompanied by the uncertain impact on vaccine effectiveness would potentially result in further waves of the epidemic. Our findings confirm that multiple measures, including public health interventions, vaccination scale-up and development of a new vaccine booster, should be integrated to counter the new challenges of new SARS-CoV-2 variants.

## Data Availability

The datasets analyzed during the current study are available in the [Johns Hopkins University Coronavirus Resource Center] repository, [https://coronavirus.jhu.edu]; [Vaccinations in the United States] repository, [https://covid.cdc.gov/covid-data-tracker/#vaccinations.]; [Variant Proportions in the US] repository, [https://covid.cdc.gov/covid-data-tracker/#variant-proportions].

## Notes

### Competing Interest Statement

The authors have declared no competing interest.

### Funding Statement

The work is supported by the Bill & Melinda Gates Foundation. LZ is supported by the National Natural Science Foundation of China (Grant number: 81950410639); Outstanding Young Scholars Funding (Grant number: 3111500001); Xi'an Jiaotong University Basic Research and Profession Grant (Grant number: xtr022019003 and xzy032020032) and Xi'an Jiaotong University Young Talent Support Grant (Grant number: YX6J004). MS was supported by the National Natural Science Foundation of China (grant number: 11801435), China Postdoctoral Science Foundation (grant number: 2018M631134, 2020T130095ZX); the Fundamental Research Funds for the Central Universities (grant number: xjh012019055); Natural Science Basic Research Program of Shaanxi Province (Grant number: 2019JQ-187); XL was supported by the Special emergency public health safety project of Shaanxi Provincial Education Department (Grant number: 20JG007).

## Reference

1. Coronavirus Resource Center: COVID-19 Tracking [https://coronavirus.jhu.edu]

2. Restrictions are easing across the UK [https://www.gov.uk/coronavirus]

3. State COVID-19 Data and Policy Actions [https://www.kff.org/report-section/state-covid-19-data-and-policy-actions-policy-actions/]

4. Israel to lift most COVID-19 restrictions in June—health minister [https://mb.com.ph/2021/05/24/israel-to-lift-most-covid-19-restrictions-in-june-health-minister/]

5. COVID-19 vaccination and blood clotting [https://www.gov.uk/government/publications/covid-19-vaccination-and-blood-clotting]

6. COVID vaccination data [https://ourworldindata.org/covid-vaccinations]

7. Plante JA, Mitchell BM, Plante KS, Debbink K, Weaver SC, Menachery VD: The variant gambit: COVID-19’s next move. Cell Host Microbe 2021, 29(4):508–515.

8. Davies NG, Abbott S, Barnard RC, Jarvis CI, Kucharski AJ, Munday JD, Pearson CAB, Russell TW, Tully DC, Washburne AD et al: Estimated transmissibility and impact of SARS-CoV-2 lineage B.1.1.7 in England. Science 2021.

9. Kow CS, Merchant HA, Hasan SS: Mortality risk in patients infected with SARS-CoV-2 of the lineage B.1.1.7 in the UK. J Infect 2021.

10. Emary KRW, Golubchik T, Aley PK, Ariani CV, Angus B, Bibi S, Blane B, Bonsall D, Cicconi P, Charlton S et al: Efficacy of ChAdOx1 nCoV-19 (AZD1222) vaccine against SARS-CoV-2 variant of concern 202012/01 (B.1.1.7): an exploratory analysis of a randomised controlled trial. The Lancet 2021.

11. Liu Y, Liu J, Xia H, Zhang X, Fontes-Garfias CR, Swanson KA, Cai H, Sarkar R, Chen W, Cutler M et al: Neutralizing Activity of BNT162b2-Elicited Serum. New England Journal of Medicine 2021.

12. Wang P, Nair MS, Liu L, Iketani S, Luo Y, Guo Y, Wang M, Yu J, Zhang B, Kwong PD et al: Antibody resistance of SARS-CoV-2 variants B.1.351 and B.1.1.7. Nature 2021.

13. Munitz A, Yechezkel M, Dickstein Y, Yamin D, Gerlic M: BNT162b2 Vaccination Effectively Prevents the Rapid Rise of SARS-CoV-2 Variant B.1.1.7 in high risk populations in Israel. Cell Rep Med 2021:100264.

14. Collier DA, De Marco A, Ferreira IATM, Meng B, Datir RP, Walls AC, Kemp SA, Bassi J, Pinto D, Silacci-Fregni C et al: Sensitivity of SARS-CoV-2 B.1.1.7 to mRNA vaccine-elicited antibodies. Nature 2021.

15. Wu K, Werner AP, Koch M, Choi A, Narayanan E, Stewart-Jones GBE, Colpitts T, Bennett H, Boyoglu-Barnum S, Shi W et al: Serum Neutralizing Activity Elicited by mRNA-1273 Vaccine. N Engl J Med 2021, 384(15):1468–1470.

16. Coronavirus variant that first appeared in India arrives in the US [https://www.usatoday.com/story/news/health/2021/05/17/coronavirus-variant-b-1-167-first-seen-india-now-us-what-know/5099593001/]

17. Polack FP, Thomas SJ, Kitchin N, Absalon J, Gurtman A, Lockhart S, Perez JL, Pérez Marc G, Moreira ED, Zerbini C et al: Safety and Efficacy of the BNT162b2 mRNA Covid-19 Vaccine. N Engl J Med 2020, 383(27):2603–2615.

18. Baden LR, El Sahly HM, Essink B, Kotloff K, Frey S, Novak R, Diemert D, Spector SA, Rouphael N, Creech CB et al: Efficacy and Safety of the mRNA-1273 SARS-CoV-2 Vaccine. N Engl J Med 2021, 384(5):403–416.

19. Sadoff J, Gray G, Vandebosch A, Cárdenas V, Shukarev G, Grinsztejn B, Goepfert PA, Truyers C, Fennema H, Spiessens B et al: Safety and Efficacy of Single-Dose Ad26.COV2.S Vaccine against Covid-19. N Engl J Med 2021.

20. COVID-19 Vaccination Program Operational Guidance [https://www.cdc.gov/vaccines/covid-19/covid19-vaccination-guidance.html#guidance-jurisdictions]

21. COVID-19 Vaccinations in the United States [https://covid.cdc.gov/covid-data-tracker/#vaccinations]

22. Tracking of Variants [https://www.gisaid.org/hcov19-variants/]

23. Variant Proportions in the US [https://covid.cdc.gov/covid-data-tracker/#variant-proportions]

24. Moghadas SM, Vilches TN, Zhang K, Wells CR, Shoukat A, Singer BH, Meyers LA, Neuzil KM, Langley JM, Fitzpatrick MC et al: The impact of vaccination on COVID-19 outbreaks in the United States. Clin Infect Dis 2021.

25. Alagoz O, Sethi AK, Patterson BW, Churpek M, Alhanaee G, Scaria E, Safdar N: The Impact of Vaccination to Control COVID-19 Burden in the United States: A Simulation Modeling Approach. medRxiv 2021.

26. Makhoul M, Chemaitelly H, Ayoub HH, Seedat S, Abu-Raddad LJ: Epidemiological Differences in the Impact of COVID-19 Vaccination in the United States and China. Vaccines (Basel) 2021, 9(3).

27. Shen M, Zu J, Fairley CK, Pagán JA, An L, Du Z, Guo Y, Rong L, Xiao Y, Zhuang G et al: Projected COVID-19 epidemic in the United States in the context of the effectiveness of a potential vaccine and implications for social distancing and face mask use. Vaccine 2021, 39(16):2295–2302.

28. Wintachai P, Prathom K: Stability analysis of SEIR model related to efficiency of vaccines for COVID-19 situation. Heliyon 2021, 7(4):e06812.

29. Sah P, Vilches TN, Moghadas SM, Fitzpatrick MC, Singer BH, Hotez PJ, Galvani AP: Accelerated vaccine rollout is imperative to mitigate highly transmissible COVID-19 variants. EClinicalMedicine 2021, 2021:100865.

30. Borchering RK, Viboud C, Howerton E, Smith CP, Truelove S, Runge MC, Reich NG, Contamin L, Levander J, Salerno J et al: Modeling of Future COVID-19 Cases, Hospitalizations, and Deaths, by Vaccination Rates and Nonpharmaceutical Intervention Scenarios - United States, April-September 2021. MMWR Morb Mortal Wkly Rep 2021, 70(19):719–724.

31. Li Q, Guan X, Wu P, Wang X, Zhou L, Tong Y, Ren R, Leung KSM, Lau EHY, Wong JY et al: Early Transmission Dynamics in Wuhan, China, of Novel Coronavirus-Infected Pneumonia. N Engl J Med 2020, 382(13):1199–1207.

32. Wang X: Safety and Efficacy of the BNT162b2 mRNA Covid-19 Vaccine. N Engl J Med 2021, 384(11).

33. Garcia-Beltran WF, Lam EC, St. Denis K, Nitido AD, Garcia ZH, Hauser BM, Feldman J, Pavlovic MN, Gregory DJ, Poznansky MC et al: Multiple SARS-CoV-2 variants escape neutralization by vaccine-induced humoral immunity. Cell 2021.

34. Muik A, Wallisch AK, Sänger B, Swanson KA, Mühl J, Chen W, Cai H, Maurus D, Sarkar R, Türeci Ö et al: Neutralization of SARS-CoV-2 lineage B.1.1.7 pseudovirus by BNT162b2 vaccine-elicited human sera. Science 2021, 371(6534):1152–1153.

35. Sapkal GN, Yadav PD, Ella R, Deshpande GR, Sahay RR, Gupta N, Mohan VK, Abraham P, Panda S, Bhargava B: Inactivated COVID-19 vaccine BBV152/COVAXIN effectively neutralizes recently emerged B 1.1.7 variant of SARS-CoV-2. J Travel Med 2021.

36. Shen X, Tang H, McDanal C, Wagh K, Fischer W, Theiler J, Yoon H, Li D, Haynes BF, Sanders KO et al: SARS-CoV-2 variant B.1.1.7 is susceptible to neutralizing antibodies elicited by ancestral spike vaccines. Cell Host Microbe 2021, 29(4):529-539.e523.

37. Abu-Raddad LJ, Chemaitelly H, Butt AA: Effectiveness of the BNT162b2 Covid-19 Vaccine against the B.1.1.7 and B.1.351 Variants. N Engl J Med 2021.

38. Haas EJ, Angulo FJ, McLaughlin JM, Anis E, Singer SR, Khan F, Brooks N, Smaja M, Mircus G, Pan K et al: Impact and effectiveness of mRNA BNT162b2 vaccine against SARS-CoV-2 infections and COVID-19 cases, hospitalisations, and deaths following a nationwide vaccination campaign in Israel: an observational study using national surveillance data. Lancet 2021, 397(10287):1819–1829.

39. Sansone E, Tiraboschi M, Sala E, Albini E, Lombardo M, Castelli F, De Palma G: Effectiveness of BNT162b2 vaccine against the B.1.1.7 variant of SARS-CoV-2 among healthcare workers in Brescia, Italy. J Infect 2021.

40. Lopez Bernal J, Andrews N, Gower C, Robertson C, Stowe J, Tessier E, Simmons R, Cottrell S, Roberts R, O’Doherty M et al: Effectiveness of the Pfizer-BioNTech and Oxford-AstraZeneca vaccines on covid-19 related symptoms, hospital admissions, and mortality in older adults in England: test negative case-control study. Bmj 2021, 2021:1088.

41. Covid booster shot generates promising immune response against variants found in South Africa, Brazil [https://www.cnbc.com/2021/05/05/covid-booster-shot-moderna-says-vaccine-generates-promising-immune-response-against-variants.html]

42. Scientist expects elderly, people with underlying conditions to be first to get Covid vaccine booster shots [https://www.cnbc.com/2021/05/04/covid-booster-shot-pfizer-expects-older-adults-those-with-underlying-conditions-to-be-first-in-line.html]

43. Boehm E, Kronig I, Neher RA, Eckerle I, Vetter P, Kaiser L: Novel SARS-CoV-2 variants: the pandemics within the pandemic. Clin Microbiol Infect 2021.

44. Cascella M, Rajnik M, Aleem A, Dulebohn SC, Di Napoli R: Features, Evaluation, and Treatment of Coronavirus (COVID-19). In: StatPearls. edn. Treasure Island (FL): StatPearls Publishing Copyright © 2021, StatPearls Publishing LLC.; 2021.

45. Researchers propose 5 strategies to encourage COVID-19 immunization [https://www.medicalnewstoday.com/articles/researchers-propose-5-strategies-to-encourage-covid-19-immunization]

