## Appendix for "Projecting the impact of SARS-CoV-2 variants on the COVID-19 epidemic and social restoration in the United States: a mathematical modelling study"

This supplementary document describes in detail the model contruction, calibration, prediction, and estimation of parameters presented in the main text.

#### 1. Model contruction

##### 1.1 Model structure and formulation

We extended a previously published dynamic compartmental model [1] to describe the circulation of two SARS-CoV-2 strains (the wildtype and B.1.1.7 variant) in the US. The population is divided into eighteen compartments (**Figure S1**), it includes susceptible individuals (S), vaccinated individuals (V), and sixteen compartments in pair infected by wild strains or B.1.1.7 variant. These sixteen compartments are latent infections (E, $E_m$), asymptomatic infections (A, $A_m$), undiagnosed infections with mild/moderate ($I_1$, $I_{m,1}$) and severe/critical symptoms ($I_2$, $I_{m,2}$), diagnosed infections with mild/moderate ($T_1$, $T_{m,1}$) and severe/critical symptoms ($T_2$, $T_{m,2}$), recovered (R, $R_m$) and deceased (D, $D_m$) cases, respectively. The total population size is denoted by N, where $N=S+V+E+A+I_1+I_2+T_1+T_2+R+E_m+A_m+I_{m,1}+I_{m,2}+T_{m,1}+T_{m,2}+R_m$. The model is described by the following system of ordinary differential equations:

$$
\begin{cases}
\dfrac{dS}{dt} = -(\lambda_{pub}+\lambda_{m,pub})(S-S_f)-(\lambda_{pri}+\lambda_{m,pri})S_f - wS, \\
\dfrac{dV}{dt} = wS - \left[(1-\varepsilon_V)\lambda_{pub}+(1-\varepsilon_{m,V})\lambda_{m,pub}\right](V-V_f) - \left[(1-\varepsilon_V)\lambda_{pri}+(1-\varepsilon_{m,V})\lambda_{m,pri}\right]V_f, \\
\dfrac{dN_f}{dt} = r\left\{(\lambda_{pub}+\lambda_{m,pub})(S-S_f)+\left[(1-\varepsilon_V)\lambda_{pub}+(1-\varepsilon_{m,V})\lambda_{m,pub}\right](V-V_f)\right\} - \xi N_f, \\
\dfrac{dS_f}{dt} = (r-1)\left\{(\lambda_{pub}+\lambda_{m,pub})(S-S_f)+\left[(1-\varepsilon_V)\lambda_{pub}+(1-\varepsilon_{m,V})\lambda_{m,pub}\right](V-V_f)\right\}\dfrac{S_f}{S_f+V_f} - (\lambda_{pri}+\lambda_{m,pri}+\xi)S_f, \\
\dfrac{dV_f}{dt} = (r-1)\left\{(\lambda_{pub}+\lambda_{m,pub})(S-S_f)+\left[(1-\varepsilon_V)\lambda_{pub}+(1-\varepsilon_{m,V})\lambda_{m,pub}\right](V-V_f)\right\}\dfrac{V_f}{S_f+V_f} - (\lambda_{pri}^V+\lambda_{m,pri}^V+\xi)V_f, \\
\dfrac{dE}{dt} = \lambda_{pub}(S-S_f)+\lambda_{pri}S_f+(1-\varepsilon_V)\lambda_{pub}(V-V_f)+(1-\varepsilon_V)\lambda_{pri}V_f - k_1E, \\
\dfrac{dE_m}{dt} = \lambda_{m,pub}(S-S_f)+\lambda_{m,pri}S_f+(1-\varepsilon_{m,V})\lambda_{m,pub}(V-V_f)+(1-\varepsilon_{m,V})\lambda_{m,pri}V_f - k_1E_m, \\
\dfrac{dA}{dt} = k_1\rho E - \gamma_0 A, \\
\dfrac{dA_m}{dt} = k_1\rho E_m - \gamma_0 A_m, \\
\dfrac{dI_1}{dt} = k_1(1-\rho)E - \alpha_1 I_1 - k_2 I_1 - \gamma_0 I_1, \\
\dfrac{dI_{m,1}}{dt} = k_1(1-\rho)E_m - \alpha_1 I_{m,1} - k_2 I_{m,1} - \gamma_0 I_{m,1}, \\
\dfrac{dI_2}{dt} = k_2 I_1 - \alpha_2 I_2 - \mu_1 I_2, \\
\dfrac{dI_{m,2}}{dt} = k_2 I_{m,1} - \alpha_2 I_{m,2} - (1+\mu_{up})\mu_1 I_{m,2}, \\
\dfrac{dT_1}{dt} = \alpha_1 I_1 - \gamma_1 T_1 - k_3 T_1, \\
\dfrac{dT_{m,1}}{dt} = \alpha_1 I_{m,1} - \gamma_1 T_{m,1} - k_3 T_{m,1}, \\
\dfrac{dT_2}{dt} = \alpha_2 I_2 + k_3 T_1 - \gamma_2 T_2 - \mu_2 T_2, \\
\dfrac{dT_{m,2}}{dt} = \alpha_2 I_{m,2} + k_3 T_{m,1} - \gamma_2 T_{m,2} - (1+\mu_{up})\mu_2 T_{m,2}, \\
\dfrac{dR}{dt} = \gamma_0 A + \gamma_0 I_1 + \gamma_1 T_1 + \gamma_2 T_2, \\
\dfrac{dR_m}{dt} = \gamma_0 A_m + \gamma_0 I_{m,1} + \gamma_1 T_{m,1} + \gamma_2 T_{m,2}, \\
\dfrac{dD}{dt} = \mu_1 I_2 + \mu_2 T_2, \\
\dfrac{dD_m}{dt} = (1+\mu_{up})\mu_1 I_{m,2} + (1+\mu_{up})\mu_2 T_{m,2}.
\end{cases}
$$

#### 1.2 The disease progression

In our model, the number of residents (*N*) are divided into households members ($N_f$) and public mebers ($N$-$N_f$) to capture varied force of infection in the public settings (e.g. public transportations, supermarkets, offices, etc) and household (home or other private settings). The number of households members ($N_f$) are dependent on the number of infected individuals and their family member (*r*), while the rest of residents denote as public mebers ($N$-$N_f$). We assumed that the number of the households at risk of infection is the same as the number of individuals infected in public settings because the probability of two or more household members being infected at the same time but at different public venues is very small. The average number of household members (*r*) in a US family was estimated as 4 [2] and the mean recovery period ($1/\xi$) of the infected family members was 28 days [3]. The details values of parameters are list in the **Table S1**.

A susceptible or vaccinated individual (S,V) may be infected by a SARS-CoV-2 strain (either the wildtype or B.1.1.7) and entered a latent infection stage (E, $E_m$, *m* denotes variant). Undergo a mean incubation of 5.2 (4.1-7.0) days, a proportion of infected individuals developed symptoms ($I_1$, $I_{m,1}$) before diagnosis and report ($T_1$, $T_{m,1}$), while the rest infected individuals (A, $A_m$) would not show any symptoms and move forward to recovery (R, $R_m$). Both undiagnosed and diagnosed infected individuals could progress to severe/critical stage ($I_2$, $I_{m,2}$, $T_2$, $T_{m,2}$) and unalterably occur to death (D, $D_m$) or recovery, but with varied progress rate on account of medical care.

#### 1.3 Force of infection

For susceptible individuals (S), the infection rates were denoted by the force of infection $\lambda_{pub}(t)$, $\lambda_{pri}(t)$, $\lambda_{m,pub}(t)$ and $\lambda_{m,pri}(t)$ as shown in eq. (1), which meant the probability of a susceptible individual infected by wildtype and B.1.1.7 variant in public settings and households. The force of infection was dependent on the number of latent, asymptomatic (E, $E_m$, A, $A_m$) and undiagnosed symptomatic infections ($I_1$, $I_{1,m}$, $I_2$, $I_{2,m}$). Previous studies had reported a 75% lower infectiveness ($\varepsilon$) of latent, asymptomatic individuals (E, A) compared with symptomatic individuals ($I_1$, $I_2$) [4]. For vaccinated individuals (V), the force of infection was supposed to decline that depends on the vaccine's effectiveness for wildtype ($\varepsilon_V$) or B.1.1.7 variant ($\varepsilon_{m,V}$) and denoted by $(1-\varepsilon_V)\lambda_{pub}(t)$, $(1-\varepsilon_V)\lambda_{pri}(t)$, $(1-\varepsilon_{m,V})\lambda_{m,pub}(t)$ and $(1-\varepsilon_{m,V})\lambda_{m,pri}(t)$.

$$\begin{aligned}
&\lambda_{pub}(t)=\left[(1-\varepsilon)(E+A)+(I_1+I_2)\right]\beta_{pub}(t)\frac{1}{N-N_f},\\
&\lambda_{pri}(t)=\left[(1-\varepsilon)(E+A)+(I_1+I_2)\right]\beta_{pri}(t)\frac{1}{N_f},\\
&\lambda_{m,pub}(t)=\left[(1-\varepsilon)(E_m+A_m)+(I_{m,1}+I_{m,2})\right](1+\beta_{up})\beta_{pub}(t)\frac{1}{N-N_f},\\
&\lambda_{m,pri}(t)=\left[(1-\varepsilon)(E_m+A_m)+(I_{m,1}+I_{m,2})\right](1+\beta_{up})\beta_{pri}(t)\frac{1}{N_f}.
\end{aligned} \tag{1}$$

Where

$$\begin{aligned}
&\beta_{pub}(t)=\beta m_{pub}(t)(1-\theta_1 p_{pub}(t))(1-\theta_2 q(t)),\\
&\beta_{pri}(t)=\beta m_{pri}(t)(1-\theta_2 q(t)),
\end{aligned} \tag{2}$$

The force of infection also depends on the effective contacts rate ($\beta_{pub}$, $\beta_{pri}$) in the population, which are the product of contact number ($m_{pub}$, $m_{pri}$) and transmission probability of per-contact with wildtype ($\beta$) or B.1.1.7 variant ($\beta_m$) infections. Previous literature has reported a 59% (56-63%) higher transmissibility in B.1.1.7 ($\beta_m$) compared with the wildtype ($\beta$) [5].

To simulate the community strategies for COVID-19 epidemic, we considered three non-pharmaceutical interventions (NPIs, i.e., restricting social distancing, insisting mask use, and keeping hand hygiene) to control the force of infection and further reduce infections and deaths.

Restricting social distancing in different epidemic stages would reduce the public contact number ($m_{pub}$) but inversely increase the household contact number ($m_{pri}$). Using masks could lower per-contact transmission probability ($\beta$, $\beta_m$) in public settings that lie with the usage percentage and effectiveness of the masking in preventing infection. Washing hands with soap and water whenever possible are also considered to prevent the spread of COVID-19 because it reduces the amount of virus on hands, which means to reduce per-contact transmission probability ($\beta$, $\beta_m$) in both public settings and household, too. In our model, the effectiveness of masking and handwashing in preventing infection were 85% (95% CI: 66-93%) and 42% (50-95%), respectively [6, 7]. Based on the trend of social interventions of public surveys [8, 9], we assumed that the three NPIs as well as the vaccination rate ($w$) change in different epidemic stages following Logistic function as follow.

$$y(t)=y_{ini}+\frac{y_{end}-y_{ini}}{1+e^{-k(t-t_{mid})}} \tag{3}$$

Logistic function is a common S-shaped function, the initial stage is roughly exponential growth or reduction; and then it slows down as it starts to saturate; finally, the increase stops when it reaches maturity. Logistic function is dependent on four parameters: (1) the initial value of growth or reduction $y_{ini}$; (2) the end value of growth or reduction $y_{end}$; (3) the rate of exponential growth or reduction $k$; (4) the time when apperence of mid value in growth or reduction $t_{mid}$. The details of Logistic function are list in the **Table S2**.

### 2. Model calibration

#### 2.1 Data sources

We collected publicly available reportable epidemiological data in the US from Johns Hopkins University Coronavirus Resource Center [10] and the Centers for Disease Control and

Prevention (CDC) [11, 12]. These websites provided daily confirmed COVID-19 infection cases and death cases from 1st March 2020 to 31st May 2021, the number of COVID-19 vaccination uptakes and variant proportions from 13th December 2020 to 8th May 2021. All four types of data were used to calibrate the model. An additional table file shows this in more detail [see **Additional file 1**]

#### 2.2 Model calibration

We calibrated the model by fitting the daily COVID-19 infections, deaths, vaccination, and proportion of variants in the US from 1$^{st}$ March 2020 to 31$^{st}$ May 2021, and predicted the daily and cumulative infections and deaths of COVID-19 over the next 12 months (from 1$^{st}$ June 2021 to 31$^{st}$ May 2022). We used the nonlinear least-squares method to minimised the Root Mean Squared Error (RMSE) between four types of model-simulated and reported data. Due to the varied orders of magnitude of four data, we standardised these data for the same orders of magnitude by dividing by the max value of the datasets, which could avoid the fitting process automatically skewing larger orders of magnitude data and better match each type of data (Figure 1). We estimated some of the model parameters by data fitting and obtained the other model parameters from the published literatures (**Table S1**).

Based on the 'calibrated' scenario, we perturbed model parameters around the 'calibrated' parameter set to generate a band of curves that best describe the data variations and retain a minimal level of RMSE. We randomly generated 200 small 'perturbating factors'. For each of the perturbing factors, we randomly sampled 100 parameter sets based on Latin Hypercube Sampling (LHS) between the parameter range generated by a random walk (adding the perturbing factor in both positive and negative directions). Hence, we obtained 200 groups of various perturbance, and each group has 100 randomised parameter sets. For each of the 200 groups, we calculated the number of data points covered by the band of curves simulated by the 100 parameter sets and their RMSE. We hence selected the one with the minimal RMSE across 200 bands as the set of simulations that best explained the observed data. The 100 curves in the selected band were used to calculate the 95% CI of the model outcomes. The vaccine effectiveness against B.1.1.7 was estimated spontaneously during model calibration. We validated that the estimated effectiveness of 88.5% produced the lowest RMSE in Figure S2.

##### 2.3 Model validation for estimated vaccines effectiveness on B.1.1.7 variant

To estimate current vaccines effectiveness on B.1.1.7 variant, we conducted the separate compartments for wildtype and B.1.1.7 to dynamically simulate total infections and deaths under the vaccine protection, which were calibrated by daily reported COVID-19 infection cases and death cases. With the known vaccine effectiveness on wildtype and its proportion of new infections, we calculated the infections and deaths caused by B.1.1.7 from total and further estimated the vaccine effectiveness on B.1.1.7.

The estimate is dependent on the model calibration and parameters optimization. To validate the reliability of estimated vaccine effectiveness, we conducted an extra comparison of the total variance (the sum of square errors of four types of data) in 0-100% vaccine effectiveness on B.1.1.7 variant (**Figure S2**). The minimum of total variance means the best match of simulations and actual epidemic data, at this point the estimated vaccine effectiveness was the best suit for available epidemic data.

#### 3. Model prediction

##### 3.1 Construction of scenarios

The emergence of E484K substitution in B.1.1.7 or other new variants may potentially reduce the effectiveness of the existing vaccines. We evaluate its impact on the COVID-19 epidemic when the vaccine effectiveness (1) adopts the model-estimated value for B.1.1.7 (baseline scenario); reduces to (2) 75%; (3) 50% and (4) 25%. For each scenario, we calculated the cumulative infections and deaths due to COVID-19 over the next 12 months (1st June 2021 to 31st May 2022). Since we did not know the viral properties of potential new variants, we also simulated nine scenarios with varying viral transmissibility and mortality as a sensitivity analysis (Figure 2).

##### 3.2 Impact of social restoration

Social restoration means releasing restrictions of daily shopping, working, meeting or gathering, and traveling, which would significantly enlarge the force of infection for both wild type and B.1.1.7 variant. To assess the timing of social restoration and herd immunity, we conducted three situations of social restoration at 60%, 65%, and 70% vaccination coverage levels and calculated the cumulative infections and deaths of COVID-19 over the next 12 months to evaluate their impact on the COVID-19 epidemic trend (Figure 3).

##### 3.3 Uncertainty and sensitivity analyses

Based on the selected 100 parameter sets in model calibration, we produced the sensitivity analysis to accommodate the uncertainty of model parameters and determine the 95% CI of the cumulative COVID-19 infections and deaths. In addition, we also explore the impact on the epidemic trends of COVID-19 in several scenarios (new variants emerge with higher transmissibility, mortality, and lower vaccine effectiveness; social restoration; Figures 2-3). All analyses and simulations were performed in MATLAB R 2019b.

**Table S1**. The values of parameters based on references or estimation by nonlinear least-squares (NLS) method in the US.

| Parameter | Description | Range or 95% CI from NLS | Source |
|---|---|---|---|
| $1/k_1$ | The mean incubation time (days) | 5.2 (4.1-7.0) | [13] |
| $1/k_2$ | The mean time from undiagnosed mild/moderate stage to undiagnosed severe/critical stage (days) | 10 | [14] |
| $k_3$ | The progression rate from diagnosed mild/moderate stage to diagnosed severe/critical stage | 0.0287 (0.0284-0.0291)$\times k_2$ | NLS |
| $1/\alpha_1$(t) | The average period from symptoms onset to diagnose for mild/moderate cases (days) | Decreased by logistic function: Mar 1, 2020-May 31, 2022: 7.74 (7.65-7.82)-3.29 (3.25-3.33) | NLS |
| $1/\alpha_2$ | The average diagnose period for severe/critical cases (days) | 2.23 (2.20-2.25) | NLS |
| $1/\gamma_0$ | The mean time for natural recovery (days) | 10.65 (10.53-10.78) | NLS |
| $1/\gamma_1$ | The average recovery period for diagnosed mild/moderate cases (days) | 7 | [15, 16] |
| $1/\gamma_2$ | The average recovery period for diagnosed severe/critical cases (days) | $21-1/\alpha_2$ | [16] |
| $r$ | The mean number of members in a family | 4 | [2] |
| $1/\xi$ | The mean recovery period for infected family members (days) | 28 | [3] |
| E(0) | The initial value of latent individuals infected by widetype | 190 (188-192) | NLS |
| $E_m(0)$ | The initial value of latent individuals infected by B.1.1.7 | 62548 (61813-63282) | NLS |
| A(0) | The initial value of asymptomatic individuals infected by widetype | 190 (188-192) | NLS |
| $I_1(0)$ | The initial value of undiagnosed mild/moderate cases infected by widetype | 190 (188-192) | NLS |
| $I_2(0)$ | The initial value of undiagnosed severe/critical cases infected by widetype | 190 (188-192) | NLS |
| $\beta$ | The per-act transmission probability in contact with infected individuals with symptoms by widetype | 0.0303 (0.0299-0.0306) | NLS |
| $\beta_m$ | The per-act transmission probability in contact with infected individuals with symptoms caused by B.1.1.7 | 59% (56-63%)$\times \beta+\beta$ | NLS and literatures [5] |
| ε | The reduction in per-act transmission probability if infection is in latent and asymptomatic stage | 75% | [4] |

| Parameter | Description | Range or 95% CI from NLS | Source |
| --- | --- | --- | --- |
| ρ | The probability that an individual is asymptomatic | 0.4964 (0.4904-0.5023) | NLS |
| $\varepsilon_V$ | The weighted effectiveness of vaccines (Pfizer, Moderna and J&J Jensen) | 91.4% (92.6%, 92.1%, 66.9%) | [17-19] |
| $\varepsilon_{m,V}$ | The effectiveness of vaccines for B.1.1.7 | 88.5% (87.4-89.5%) | NLS |
| $\theta_1$ | The effectiveness of mask in preventing infection | 0.85 (0.66-0.93) | [6] |
| $\theta_2$ | The effectiveness of handwashing in preventing infection | 0.42 (0.1-0.95) | [7] |
| $\mu_1(\mathrm{t})$ | Disease-induced death rate of undiagnosed severe/critical cases infected by widetype | Decreased by logistic function: Mar 1, 2020-May 31, 2022: 0.0527 (0.0521-0.0533)-0.0224 (0.0222-0.0226) | NLS |
| $\mu_2(t)$ | Disease-induced death rate of diagnosed severe/critical cases infected by widetype | 0.0358 (0.0354-0.0363)× $\mu_1(\mathrm{t})$ | NLS |
| $\mu_{\mathrm{m},1}$ (t) | Disease-induced death rate of undiagnosed severe/critical cases infected by B.1.1.7 | 45% (18-78%)× $\mu_1(\mathrm{t}) + \mu_1(\mathrm{t})$ | NLS and literatures [20] |
| $\mu_{\mathrm{m},2}(t)$ | Disease-induced death rate of diagnosed severe/critical cases infected by B.1.1.7 | 0.0358 (0.0354-0.0363)× $\mu_{\mathrm{m},1}(\mathrm{t})$ | NLS and literatures [20] |
| $m_{pub,ini}$ | Base daily contact number in the public settings | 34.81 (34.41-35.20) | NLS |
| $m_{pri,ini}$ | Base daily contact number in the households | 4 | [21] |
| $m_{pub}(t)$ | The percentage of base daily contact number in the public settings | Shown in the Table S2 | NLS and literatures [8, 9] |
| $m_{pri}(t)$ | The percentage of base daily contact number in the households | Shown in the Table S2 | NLS and literatures [8, 9] |
| $p_1(t)$ | The usage percentage of face mask in the public settings | Shown in the Table S2 | NLS and literatures [8] |
| $q(t)$ | The usage percentage of handwashing | Shown in the Table S2 | NLS and literatures [22, 23] |
| $w(t)$ | The vaccination rate for susceptible individuals (per day) | Shown in the Table S2 | NLS |

**Table S2**. The details of Logistic function based on trend of social interventions of public surveys or estimation by nonlinear least-squares (NLS) method.

| parameters | | Mar 1 2020- Jun 4, 2020 | Mar 1 2020- Sep 5, 2020 | Mar 1, 2020- Mar 31, 2022 | Jun 5 2020- Sep 5 2020 | Sep 6 2020- Dec 17, 2020 | Dec 18 2020- Mar 3, 2021 | Mar 4, 2021- Apr 2, 2021 | Apr 3, 2021- May31, 2022 |
|---|---|---|---|---|---|---|---|---|---|
| $m_{pub}(t)$ | Initial | 100.00% | — | — | 16.8% | 45.2% | 66.8% | 62.5% | 83.5% |
| | end value | 16.8% | — | — | 45.2% | 66.8% | 62.5% | 83.5% | 67.3% |
| | rate | 0.2159 | — | — | 0.2159 | 0.1699 | 0.1699 | 0.2159 | 0.2159 |
| | mid-point | Api 2, 2020 | — | — | Jun 4, 2020 | Sep 5, 2020 | Dec 17, 2020 | Mar 3, 2021 | Apr 2, 2021 |
| $m_{pri}(t)$ | Initial | 100.00% | — | — | 200.00% | 165.9% | 139.9% | 145.1% | 119.9% |
| | end value | 200.00% | — | — | 165.9% | 139.9% | 145.1% | 119.9% | 139.3% |
| | rate | 0.2159 | — | — | 0.2159 | 0.1699 | 0.1699 | 0.2159 | 0.2159 |
| | mid-point | Api 2, 2020 | — | — | Jun 4, 2020 | Sep 5, 2020 | Dec 17, 2020 | Mar 3, 2021 | Apr 2, 2021 |
| $p_{pub}(t)$ | Initial | — | 0.00% | — | — | 51.4% | 45.9% | 64.3% | 46.9% |
| | end value | — | 51.4% | — | — | 45.9% | 64.3% | 46.9% | 62.4% |
| | rate | — | 0.2159 | — | — | 0.1699 | 0.1699 | 0.2159 | 0.2159 |
| | mid-point | — | Jul 12, 2020 | — | — | Sep 5, 2020 | Dec 17, 2020 | Mar 3, 2021 | Apr 2, 2021 |
| $q(t)$ | Initial | — | — | 77% | — | — | — | | — |
| | end value | — | — | 95% | — | — | — | | — |
| | rate | — | — | 0.2159 | — | — | — | | — |
| | mid-point | — | — | Api 2, 2020 | — | — | — | | — |
| $w(t)$ | Initial | — | — | 0 | — | — | — | | — |
| | end value | — | — | 0.0188 | — | — | — | | — |
| | rate | — | — | 0.0215 | — | — | — | | — |
| | mid-point | — | — | Apr 10, 2021 | — | — | — | | — |

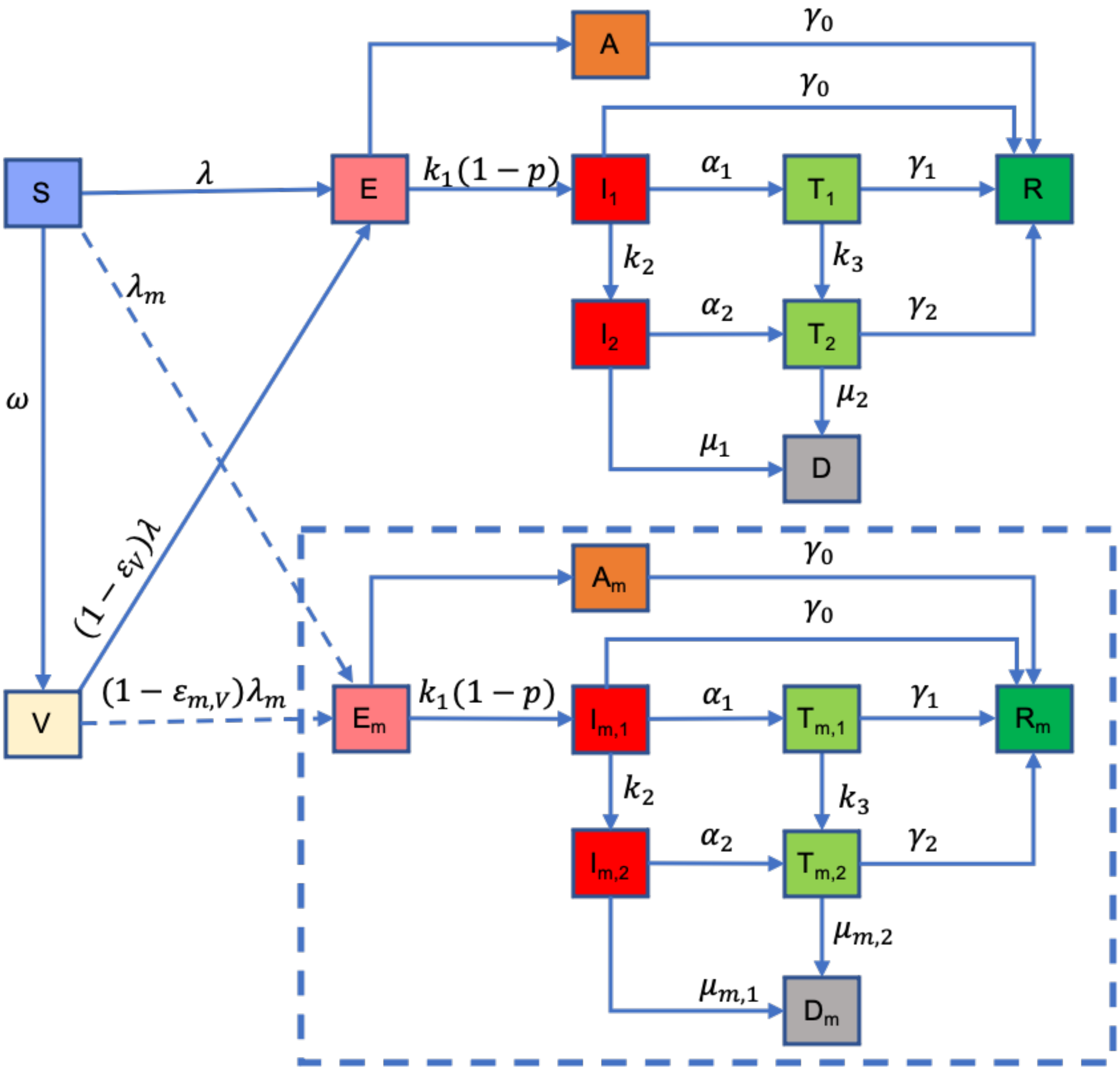


**Figure S1**. A schematic flow diagram of the transmission of COVID-19 and its mutants.

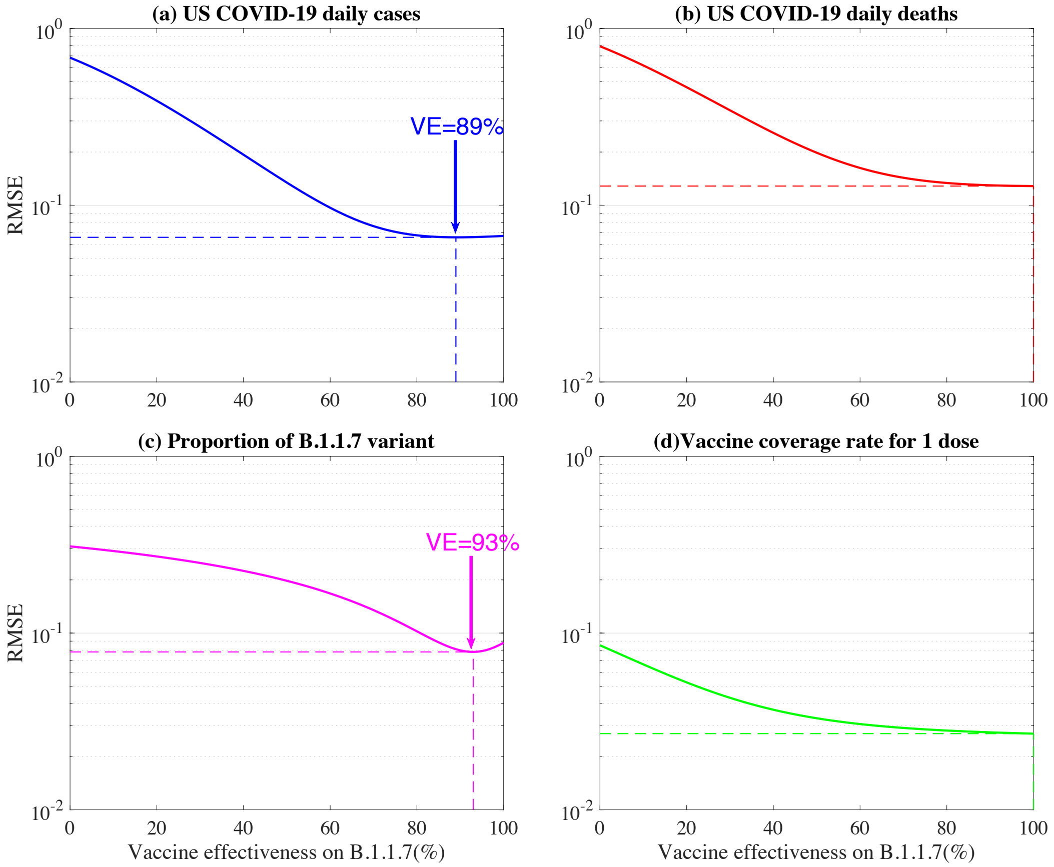


**Figure S2.** The validation for estimated vaccine effectiveness on B.1.1.7 variant. The four colored line denote the average standardized variance between four types of simulated and reported data in 0-100% vaccine effectiveness on B.1.1.7 variant. The arrows point to the lowest point of the simulated curve.

#### Supplement Reference


1. Shen M, Zu J, Fairley CK, Pagán JA, An L, Du Z, Guo Y, Rong L, Xiao Y, Zhuang G *et al*: **Projected COVID-19 epidemic in the United States in the context of the effectiveness of a potential vaccine and implications for social distancing and face mask use**. *Vaccine* 2021, **39**(16):2295-2302.
2. **Average size of households in the U.S. 2019. Statista.** [https://www.statista.com/statistics/183648/average-size-of-households-in-the-us/]
3. Shen M, Peng Z, Guo Y, Rong L, Li Y, Xiao Y, Zhuang G, Zhang L: **Assessing the effects of metropolitan-wide quarantine on the spread of COVID-19 in public space and households**. *Int J Infect Dis* 2020, **96**:503-505.
4. Prem K, Liu Y, Russell TW, Kucharski AJ, Eggo RM, Davies N, Jit M, Klepac P: **The effect of control strategies to reduce social mixing on outcomes of the COVID-19 epidemic in Wuhan, China: a modelling study**. *Lancet Public Health* 2020, **5**(5):e261-e270.
5. Davies NG, Abbott S, Barnard RC, Jarvis CI, Kucharski AJ, Munday JD, Pearson CAB, Russell TW, Tully DC, Washburne AD *et al*: **Estimated transmissibility and impact of SARS-CoV-2 lineage B.1.1.7 in England**. *Science* 2021.
6. Chu DK, Akl EA, Duda S, Solo K, Yaacoub S, Schünemann HJ: **Physical distancing, face masks, and eye protection to prevent person-to-person transmission of SARS-CoV-2 and COVID-19: a systematic review and meta-analysis**. *Lancet* 2020, **395**(10242):1973-1987.
7. Fung IC, Cairncross S: **Effectiveness of handwashing in preventing SARS: a review**. *Trop Med Int Health* 2006, **11**(11):1749-1758.
8. **Delphi survey results about the spread and impact of the COVID-19 pandemic in the United States** [https://delphi.cmu.edu/covidcast/survey-results/?region=9&date=20210511]
9. **Understand the impact of COVID-19 using real data** [https://www.unacast.com/covid19/social-distancing-scoreboard]
10. **Coronavirus Resource Center: COVID-19 Tracking** [https://coronavirus.jhu.edu]
11. **COVID-19 Vaccinations in the United States** [https://covid.cdc.gov/covid-data-tracker/#vaccinations]
12. **Variant Proportions in the US** [https://covid.cdc.gov/covid-data-tracker/#variant-proportions]
13. Li Q, Guan X, Wu P, Wang X, Zhou L, Tong Y, Ren R, Leung KSM, Lau EHY, Wong JY *et al*: **Early Transmission Dynamics in Wuhan, China, of Novel Coronavirus-Infected Pneumonia**. *N Engl J Med* 2020, **382**(13):1199-1207.
14. Wang D, Hu B, Hu C, Zhu F, Liu X, Zhang J, Wang B, Xiang H, Cheng Z, Xiong Y *et al*: **Clinical Characteristics of 138 Hospitalized Patients With 2019 Novel Coronavirus-Infected Pneumonia in Wuhan, China**. *Jama* 2020, **323**(11):1061-1069.
15. Huang C, Wang Y, Li X, Ren L, Zhao J, Hu Y, Zhang L, Fan G, Xu J, Gu X *et al*: **Clinical features of patients infected with 2019 novel coronavirus in Wuhan, China**. *Lancet* 2020, **395**(10223):497-506.
16. **Report of the WHO-China Joint Mission on Coronavirus Disease 2019 (COVID-19)** [https://www.who.int/publications/i/item/report-of-the-who-china-joint-mission-on-coronavirus-disease-2019-(covid-19)]
17. Baden LR, El Sahly HM, Essink B, Kotloff K, Frey S, Novak R, Diemert D, Spector SA, Rouphael N, Creech CB *et al*: **Efficacy and Safety of the mRNA-1273 SARS-CoV-2 Vaccine**. *N Engl J Med* 2021, **384**(5):403-416.
18. Polack FP, Thomas SJ, Kitchin N, Absalon J, Gurtman A, Lockhart S, Perez JL, Pérez Marc G, Moreira ED, Zerbini C *et al*: **Safety and Efficacy of the BNT162b2 mRNA Covid-19 Vaccine**. *N Engl J Med* 2020, **383**(27):2603-2615.
19. Sadoff J, Gray G, Vandebosch A, Cárdenas V, Shukarev G, Grinsztejn B, Goepfert PA, Truyers C, Fennema H, Spiessens B *et al*: **Safety and Efficacy of Single-Dose Ad26.COV2.S Vaccine against Covid-19**. *N Engl J Med* 2021.
20. Kow CS, Merchant HA, Hasan SS: **Mortality risk in patients infected with SARS-CoV-2 of the lineage B.1.1.7 in the UK**. *J Infect* 2021.
21. Ferguson NM, Cummings DA, Fraser C, Cajka JC, Cooley PC, Burke DS: **Strategies for mitigating an influenza pandemic**. *Nature* 2006, **442**(7101):448-452.
22. Park CL, Russell BS, Fendrich M, Finkelstein-Fox L, Hutchison M, Becker J: **Americans' COVID-19 Stress, Coping, and Adherence to CDC Guidelines**. *J Gen Intern Med* 2020, **35**(8):2296-2303.
23. Pogrebna G, Kharlamov A: **The Impact of Cross-Cultural Differences in Handwashing Patterns on the COVID-19 Outbreak Magnitude**; 2020.
